# Asynchronous influenza vaccination and adverse maternal-child health outcomes in the Brazilian semiarid, 2013 to 2018: the INFLUEN-SA Study

**DOI:** 10.1101/2020.08.24.20180455

**Authors:** José Q Filho, Francisco S Junior, Thaisy BR Lima, Vânia AF Viana, Jaqueline SV Burgoa, Alberto M Soares, Álvaro M Leite, Simone A Herron, Hunter L Newland, Kunaal S Sarnaik, Gabriel F Hanson, Jason A Papin, Sean R Moore, Aldo AM Lima

**Author notes:** R. Cel. Nunes de Melo, 1315, Rodolfo Teófilo, Fortaleza, Ceará, Brasil, CEP 60.430-270. Phones: 55 (85) 3366-8445 or −8437.

## Abstract

Recent models indicate seasonal influenza transmission in Brazil begins annually in the semiarid state of Ceará (pop. 8.8M)—before vaccine campaigns begin. To assess the extent and maternal-child health consequences of this misalignment, we tracked severe acute respiratory infections (SARI), influenza, and influenza immunizations from 2013-2018. Of 3,297 SARI cases, 145 (4%) occurred in pregnancy. Vaccine coverage was >80%; however, campaigns often occurred during or after peak influenza. Birth weights nadired and prematurity increased 30-40 weeks following peak influenza, by a magnitude of 40g and 10.7% to 15.5%, respectively. We identified 61 babies of mothers with gestational SARI; they weighed 10% less at birth (P = 0·019) and were more often premature (OR: 2.944; 95% CI: 1.100 – 7.879) relative to controls (n=122). Mistiming of influenza vaccination adversely impacts pregnancy and birth outcomes in Ceará, with critical implications for influenza transmission dynamics nationally.

## Introduction

Respiratory infections are a leading cause of morbidity and mortality worldwide (*1,2*). Complicated respiratory infections are well-recognized as an important cause of death in children and older adults. However, the impact of respiratory infections on pregnant women and fetal development are understudied, particularly in regards to the burden of disease in low- and middle-income countries and adverse birth outcomes, early childhood growth, and neurodevelopment.

Influenza epidemics are associated with high morbidity and mortality, usually in the form of excess rates of pneumonia, associated hospitalizations, and deaths (*3*). Pregnant women and their infants are considered a high risk group for severe influenza disease (*4,5*). Recently, Regan et al. conducted a retrospective cohort study of pregnant women from Australia, Canada, Israel, and the United States and showed that acute respiratory or febrile illness (ARFI) hospitalizations with and without influenza were associated with low birthweight, but not small-for-gestational-age *(6)*. A prospective cohort study of pregnant women in India, Peru and Thailand demonstrated influenza in pregnancy is associated with late pregnancy loss and reductions in mean birth weight *(7)*. During the 2009 pandemic, meta-analysis showed the risk of hospitalization for influenza was two-fold higher during pregnancy (*8*), with potential consequences for the physical and neurocognitive development of children born to mothers infected during pregnancy. These consequences were similar to the effects reported in studies related to maternal and child undernutrition as well as early childhood undernutrition associated with diarrheal diseases resulting in impaired physical fitness and cognitive function 4-7 years later in rural and urban communities (*9-13*).

In Brazil, recent time-series analysis by Almeida and colleagues revealed 12 of 27 states demonstrate annual-seasonal influenza activity (*14)*. This seasonal influenza is clustered along the coast, whereas Amazonian and Central West states exhibit no readily identifiable seasonality, likely due to more complex and difficult to detect seasonal patterns of influenza spread. Peak seasonal influenza activity begins in the semiarid state of Ceará (population 8.8 million) typically in mid-May, before subsequent transmission southward (*4*). However, influenza circulation begins as early as mid-March. Prior single center studies from the state capital of Fortaleza (population 2.6 million) showed seasonal influenza peaks 2 to 3 months earlier than in the South and Southeast of Brazil (*15*). Despite these well-described epidemiological differences, all of Brazil is subject to the same vaccination schedule, which in the semiarid region typically coincides with or follows peak influenza activity **(Figure 1)**. Because vaccine-acquired immunity against influenza typically occurs two weeks post-immunization, we hypothesized pregnant women and their fetuses in the semiarid region of Brazil may be inadequately protected against influenza.

**Figure 1.**
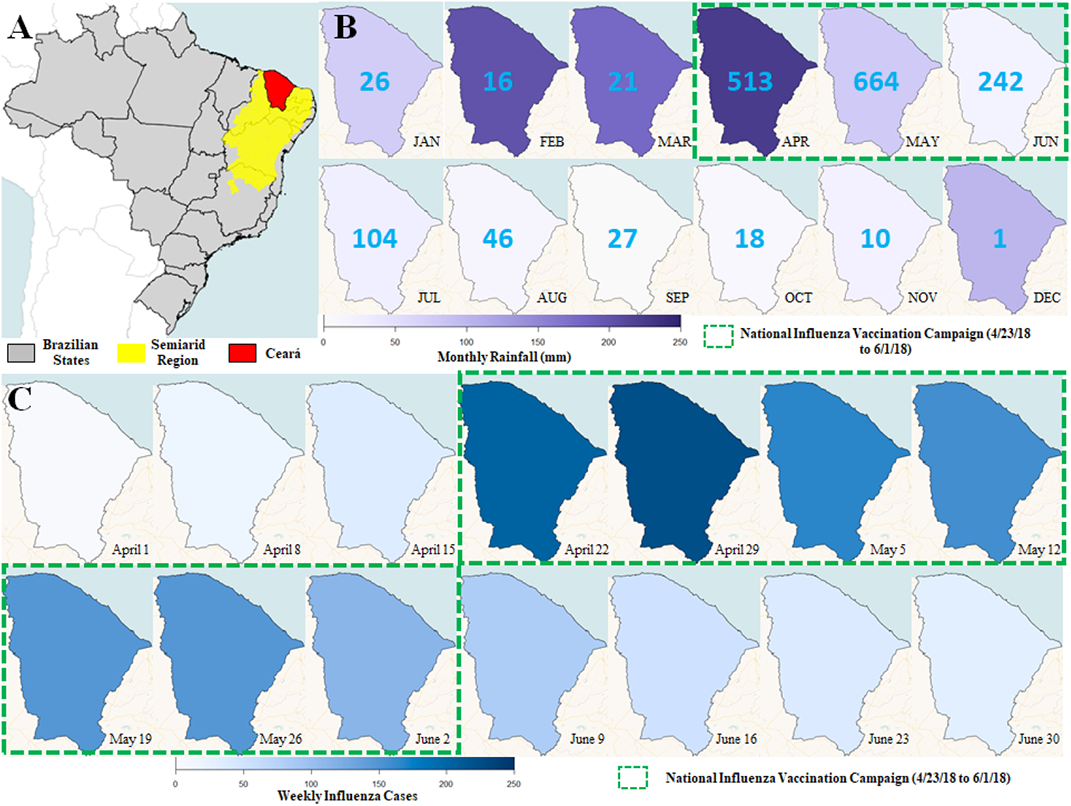
Map of Ceará and visualizations of average monthly rainfall and weekly influenza cases in Ceará, 2018. **(A)** Ceará (red) in relation to Brazil’s semiarid region (yellow) and other Brazilian states (light gray). **(B)** Visualizations of average monthly rainfall (mm) across the state of Ceará over the course of 2018 with total monthly influenza cases labeled in the center of each visualization. Brazil’s National Influenza Vaccination Campaign (green bounded box) was conducted from April 23, 2018 to June 1, 2018. Average monthly rainfall steadily increased from January to April and then decreased from April to September before increasing again at the end of the year. Monthly influenza cases drastically increased from March to April, peaking in May at 664 before decreasing throughout the remainder of the year. **(C)** Visualizations of weekly influenza cases before, during, and after Brazil’s National Influenza Vaccination Campaign (green bounded box). Weekly cases quickly peaked two weeks into the campaign before beginning to steadily decrease throughout the remainder of the campaign and shortly after.

Therefore, our study had three objectives: (1) determine the extent to which pregnancy-associated SARI correlates with statewide trends in low birthweight and premature birth in Ceará; (2) evaluate timing of national influenza vaccine campaigns relative to statewide patterns of SARI and influenza; and (3) determine if documented cases of pregnancy-associated SARI are correlated with low birth weight and prematurity, after adjusting for known clinical confounders.

## Methods

### Ethical approval

We conducted this study with approval from the Ethical Review Committees of the Federal University of Ceará, Fortaleza, CE, Brazil (register) and the State Health Secretariat, Fortaleza, CE, Brazil (register at CGTES/NUVEN). This study used guaranteed public access information under the terms of Law # 12,527, of November 18, 2011, and used aggregated information from deidentified databases in a manner consistent with the provisions of resolution CNS # 510, April 7, 2016 (Official Gazette of the Union; Published on: 24/05/2016, Edition: 98, Section: 1, Page: 44, National Health Council, Ministry of Health, Brasília, Brazil).

### Study design and population

We identified cases of severe acute respiratory infection (SARI) cases registered in the state of Ceará, Brazil from 2013 to 2018 using the Notifiable Diseases Information System (SINAN-Influenza). A case of severe acute respiratory infection was defined as an individual with fever, even if referred, accompanied by cough, sore throat, dyspnea, O2 saturation <95% or respiratory discomfort, with onset of symptoms in the preceding seven days. In previous studies, multivariate regression analysis has shown cough and fever are the best predictors of laboratory-confirmed influenza (*16*). From the SINAM-Influenza case report form we collected data on demographics, education, clinical signs and symptoms, epidemiological risk factors, vaccination status, treatments received, samples collected (nasopharyngeal secretions, bronchial aspirations, tissue or others) and RT-PCR laboratory results for influenza.

### Molecular detection of influenza

RT-PCR detection of influenza viruses was based on a protocol frequently updated and published by WHO’s Global Influenza Surveillance Network (*17*). The RT-PCR assay was developed to detect seasonal influenza A, B, H1, H3 and avian H5 serotypes. Samples from the deep nostrils (nasal swab), throat (oropharyngeal swab) and nasopharynx (nasopharyngeal swab) were taken from patients and nucleic acid was extracted. The QIAamp Viral Mini Kit (Qiagen) was used for sample extraction following the manufacturer’s recommended protocols. Extracted viral RNA was subjected to a series of reverse-transcription polymerase chain reactions (RT-PCR) allowing template viral RNA to be reverse transcribed, producing complementary DNA (cDNA), which was then amplified and detected. Protocols for the RT-PCR detection and subtyping are published with detailed laboratory information by WHO’s Global Influenza Surveillance Network (*15*). The RT-PCR is a rapid and sensitive method for detection of influenza viruses and targeted genes including the type A influenza matrix gene and the haemagglutinin gene for influenza B and A subtypes: A(H1N1)pdm09 virus, A(H3N2), and former seasonal A(H1N1).

### Detection of severe acute respiratory infections during pregnancy and linkage to birth data

We constructed a database by linking information from the SINAN-Influenza database with data from the declaration of live birth certificates (SINASC database). Due to the amount of records (517,347 records) in the SINASC database, MySQL database software version 5.0.11 (Oracle Corporation, Redwood City, CA), R statistical package 3.6.2 with ’genderBR’ package 1.1.0, and Stata 11 software (Stata Corporation, College Station, TX) were used to construct and manage the combined database. This combined database was joined into a single table containing information on severe acute respiratory infection cases and birth records from pregnant women with documented severe acute respiratory infection in the SINAN-Influenza database. Separately, we linked deidentified data from individual pregnant women with and without SARI to birth certificate data for a case-control study. Where available, we linked influenza test results to these records, however sampling was limited. From birth certificate data, we collected children’s birth weight, mother’s educational level, Apgar scores (Appearance, Pulse, Grimace, Activity, Respiration), and information concerning demographics, previous and current pregnancies, and mode of delivery.

### Maternal-fetal effects of severe acute respiratory infection and design of observational descriptive study

In order to evaluate the harmful maternal-fetal effects of severe acute respiratory infection on birth weight and gestational length, we designed an observational descriptive study in which one group was defined as children born to mothers who experienced a severe acute respiratory infection during pregnancy. The second, control group was made up of twice as many randomly selected births from children born to mothers matched by age (within a range of three months) with no record of severe acute respiratory infection during the pregnancy. Birth weights were collected using SINAM influenza case records. Thus, birth weight methodology rigor was from routine clinical practice. Birth weight data from case and controls were obtained from the SINAN-Influenza database that collects data from routine clinical practices.

### Annual periodicity in birth weight and gestational length

Due to seasonal increases in influenza infection and their effect on birth outcomes, we investigated the periodicity associated with birth weight and length of gestation in Ceará. Birth outcomes were compiled from the SINASC database, where gestational length is classified with scores ranging from 1 to 6: 1, <22 weeks; 2, 22-27 weeks; 3, 28-31 weeks; 4, 32-36 weeks; 5, 37-41 weeks; and 6, post-term >42 weeks gestation (see Table 4). Preterm birth was defined as a gestational length <37 weeks. Birth weights and length of gestation was averaged by epidemiological week between 2013 and 2018 and plotted as discrete time points.

### Sample size and statistical analysis

We estimated the sample size needed to detect an effect of SARI on birth weights to be 183 total children. A sample size of 61 cases and 122 controls provided a statistical power of 80% at *P* <0·05 for children who were 10% underweight compared to controls (*17,18*). The formulae used for sample size determination to compare the two children’s birth weight means (sample size of each group) was the following: n1 = (*u*+*v*)^2^(σ_1_ ^2^+σ_2_ ^2^/*K*)/(µ_1_ -µ_2_)^2^; where µ_1_-µ_2_ = Difference between the means; σ_1_, σ_2_= Standard deviations; *u*= One-side percentage point of the normal distribution corresponding to 100%-the power, e.g. if power=80%, *u*=0·84; *v*= Percentage point of the normal distribution corresponding to the (two-side) significance level, e.g. if significance level=5%, *v*=1.96; *K*=n_2_/n_1_. For sample size calculations we used ClinCalc.com (https://clincalc.com/Stats/SampleSize.aspx).

Data were entered into spreadsheets and checked by two independent researchers to ensure accuracy. Statistical analysis was performed using SPSS (Version 20·0, IBM Corporation, Armonk, NY) (*22*). All data were analyzed anonymously. The Shapiro-Wilk test was used to evaluate normality of quantitative data. The Levene test was used to evaluate equality of variances. The Mann-Whitney test (two groups) was used for nonparametric variables. Qualitative variables were analyzed using the chi-square test or Fisher’s test. GraphPad Prism software (3.0 for Windows (San Diego, CA, USA)) was used for complementary statistical analysis, table formatting and figures. Adjusted and non-adjusted multivariate logistic regression models were used to assess underweight and preterm birth associations. Some data were coadjusted to ensure accuracy of final analyses without possible negative influence from confounding variables. Co-adjusted variables included sex, mother schooling data, information from previous and current pregnancies, and delivery information. Odds ratios or relative risk ratios with 95% confidence intervals (95% CI) were utilized to assess the risk between a variable and its outcome. All statistical tests were two-sided with a significance level of <0·05.

## Results

Our study strategy is summarized in **Figure 2**. Using the SINAM database, we identified a total of 3,298 cases of SARI from 2013 to 2018 in Ceará (**Table 1**). From these we identified 145 pregnant women with SARI. We then linked the SINAM and SINASC (live birth certificates) databases to identify children born to mothers who experienced at least one episode of severe acute respiratory infection during pregnancy. By linking these databases, we were able to identify 61 children we classified as cases. Using these same databases, we identified control children born to age-matched pregnant women with no history of reported severe acute respiratory infections during their pregnancy. We identified 122 control children, twice as many as the number of case children born to mothers who had severe acute respiratory infection during pregnancy, for a total of 183 children. SARI detection was captured using the national single surveillance system for severe acute respiratory infection using the SINAM-influenza record. Both public and private hospitals are required to report this disease to the Ministry of Health for coordinating epidemics, vaccine development and scheduling influenza vaccinations.

**Table 1.** Population characteristics, influenza and subtypes, incidence, deaths, lethality and vaccination coverage of the historical series of cases of severe acute respiratory infection, state of Ceará, Brazil - 2013 to 2018.

| Variables | Years of Monitoring |  |  |  |  |  |
| --- | --- | --- | --- | --- | --- | --- |
| Year | 2013 | 2014 | 2015 | 2016 | 2017 | 2018 |
| **Severe acute respiratory infection notified** | 330 | 173 | 289 | 546 | 285 | 1674 |
| **Male (N /%)** | 141<br>(43) | 68<br>(39) | 115<br>(40) | 272<br>(50) | 150<br>(53) | 854<br>(51) |
| **Age (median, variation)** | 26.02<br>(0-96) | 26.54<br>(0-97) | 23.11<br>(0-94) | 16.85<br>(0-100) | 1.58<br>(0-94) | 5.26<br>(0-102) |
| **Children <6 months (N /%)** | 47<br>(14) | 46<br>(27) | 76 (26) | 52 (10) | 88<br>(31) | 235<br>(14) |
| **Children 6 months to 5 years (N /%)** | 43<br>(13) | 16 (9) | 23 (8) | 174<br>(32) | 84<br>(30) | 594<br>(35) |
| **Older Adults ≥60 years (N /%)** | 53<br>(16) | 27<br>(16) | 30 (10) | 102<br>(19) | 21 (7) | 247<br>(32) |
| **Pregnant women (N /%)** | 38<br>(12) | 13 (8) | 32 (11) | 17 (3) | 10 (4) | 35 (2) |
| **Severe acute respiratory infection etiology**<br>**N (%)** | 330 | 173 | 289 | 546 | 285 | 1674 |
| Influenza | 56<br>(17) | 24<br>(14) | 55 (19) | 107<br>(20) | 36<br>(13) | 451<br>(27) |
| Other viruses / etiologic agents | 61<br>(18) | 22<br>(13) | 36 (12) | 64 (12) | 101<br>(35) | 21 (1) |
| Unspecified | 213<br>(65) | 127<br>(73) | 198<br>(69) | 375<br>(69) | 148<br>(52) | 1202<br>(72) |
| **Incidence of severe acute respiratory infection due**<br>**to influenza laboratory certificate (%)** | 17.0 | 13.9 | 20.1 | 19.6 | 12.3 | 26.9 |
| **Influenza subtypes:** | 56<br>(100) | 24<br>(100) | 58<br>(100) | 107<br>(100) | 35<br>(100) | 450<br>(100) |
| *A H1N1* (N / %) | 30<br>(54) | 18<br>(75) | 1 (2) | 89 (83) | 2 (6) | 309<br>(69) |
| *A H1 / seasonal* (N / %) | - | - | 45 (78) | - | 1 (3) | 0(0) |
| *A H3 / seasonal* (N / %) | 2 (4) | - | - | - | 25 (71) | 23 (5) |
| *A without subtype* (N / %) | 22 (39) | 1 (4) | 4 (7) | 16 (15) | - | 14 (3) |
| *B* (N / %) | 2 (4) | 5 (21) | 8 (14) | 2 (2) | 7 (20) | 104 (23) |
| **Deaths with severe acute respiratory infection** (N / %) | 13 | 2 | 1 | 40 | 24 | 159 |
| Deaths due to influenza | 9 (69) | 1 (50) | 0 (0) | 17 (43) | 5 (21) | 75 (47) |
| Deaths due to other viruses / etiologic agents / unspecified | 4 (31) | 1 (50) | 1(100) | 23 (58) | 19 (79) | 84 (53) |
| **Lethality of severe acute respiratory infection by influenza laboratory certificate** |  |  |  |  |  |  |
| Lethality - influenza (%) | 16.1 | 4.2 | 0 | 15.9 | 20.8 | 16.6 |
| **Influenza vaccine coverage, Ceará <sup>a</sup>** | 88 | 84 | 83 | 91 | 90 | - |

**Source:** Secretary of Health of the State of Ceará; Sinam case report form for severe acute respiratory infection.
<sup>a</sup> %; risk population estimated at 2.6 million. Description of risk populations: Children ages 6 months to under 6 years (5 years, 11 months and 29 days); Individuals aged 60 or over; Pregnant women; Postpartum women (up to 45 days after delivery); healthcare workers; teachers; indigenous people; carriers of chronic non-communicable diseases and other special clinical conditions; Adolescents and young people from 12 to 21 years of age living in poverty; prisoners; prison system officials; military police, civilians, firefighters and armed forces.

**Figure 2.**
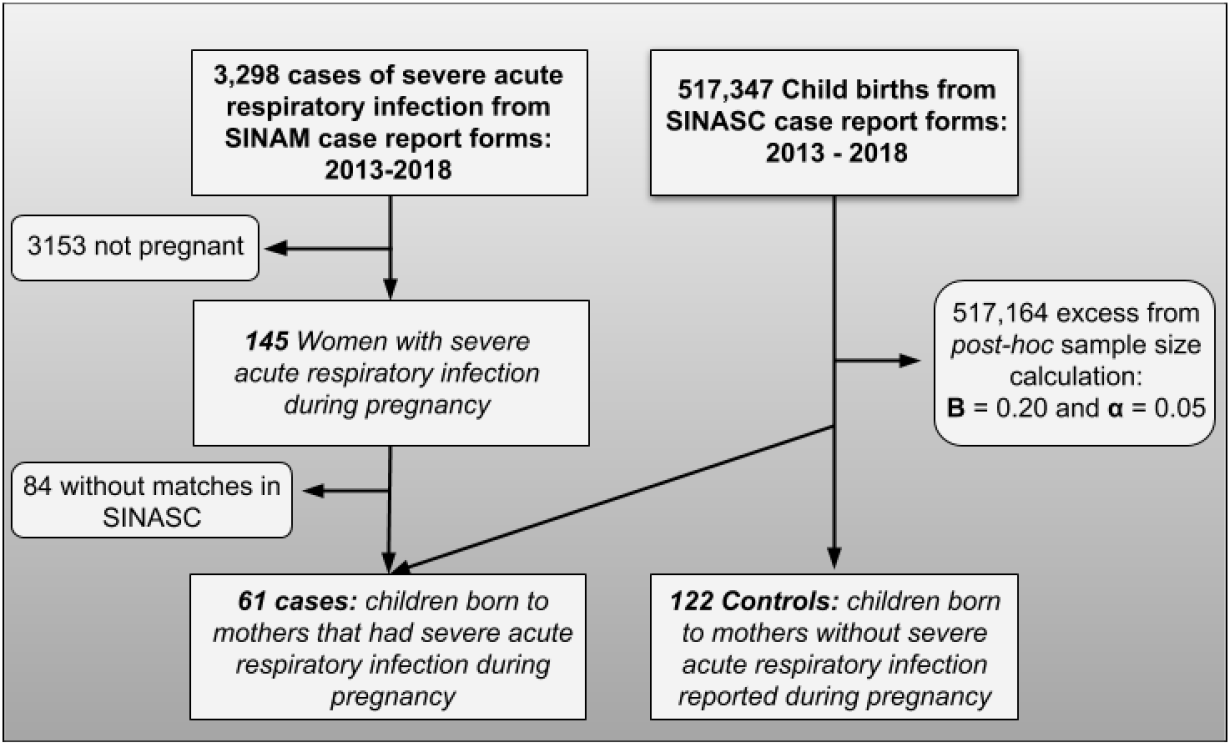
Flowchart of cases with severe acute respiratory infection, pregnant women with severe acute respiratory infection and birth outcomes in Ceará, Brazil, 2013-2018. Using SINAM data, we identified 3,298 documented cases of severe acute respiratory infection, of which 145 were in pregnant women. From this pregnant cohort, we identified 61 liveborn children by crossing SINAM and SINASC database case report forms over the same period. Of the 517,347 liveborn children, we identified 122 children born to women with no reported severe acute respiratory infections and matched these children by maternal age to create the control group.

Population characteristics are summarized in **Table 1** by sex, age, and high risk status (children, older adults, or pregnant) from 2013-2018. We observed equal proportions of cases in males and females. In general, we found more cases in children under five, ranging from 27-62% of the affected population, over the study period, with an exceptionally high burden in children younger than 6 months, ranging from 10% to 31% of the total cases. The next most abundant populations were older adults (7-32%) and pregnant women (2-38%). **Figure 3A** shows identified subtypes of influenza virus. We noted cases of influenza A/sH1N1 throughout the study period, predominantly in 2013-2014 (54% and 75%, respectively), followed by the years 2016 (83%) and 2018 (69%). In 2018, we observed the highest number of cases of SARI, associated with influenza A/H1N1, followed by influenza B (23%). We observed sporadic cases of seasonal influenza A/H1 in 2015 and 2017 and seasonal influenza A/H3 in the years 2013 and 2017-2018 (**Table 1** and **Figure 3B**). Lethality due to influenza varied from 0-21%, with the period of greatest lethality (21%) associated with a predominance of the seasonal influenza A/H3 subtype.

**Figure 3.**
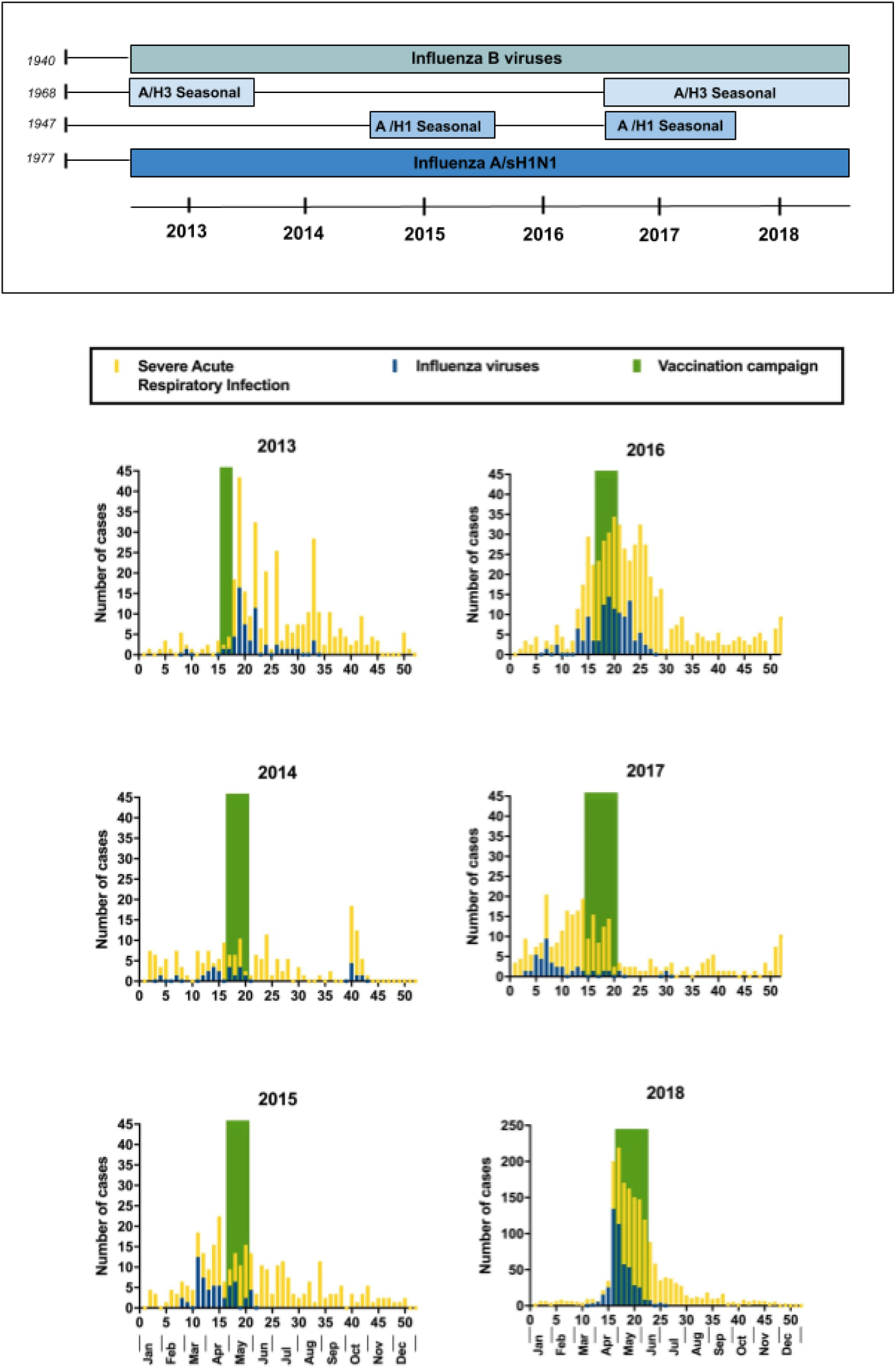
Annual circulating of influenza virus subtypes in Ceará, Brazil, 2013-2018. **(A)** The year of the origins of pandemic viruses is shown together with the duration of the influenza B virus throughout the study period as well as the influenza A/sH1N1 virus. The seasonal influenza A/H3 virus appeared in 2013 and between 2017-2018. The seasonal influenza A/H1 virus appeared in 2015 and 2017. **(B) Number of severe acute respiratory infection cases, cases of influenza viruses documented by laboratory RT-PCR and period of influenza vaccination**. The total number of cases of severe acute respiratory infection and the consistency in the increase in documented cases of influenza viruses and severe acute respiratory infection was variable over seasons and years. Alarmingly, influenza vaccination campaigns either coincided (2013, 2014 and 2016) or immediately followed peak disease activity (2015, 2017, and 2018).

**Table 2** summarizes characteristics of pregnant women with SARI. Their median age was 26 years old with range from 15 to 44 years old. Of 145 total pregnant cases with SARI, 43 (32%) had laboratory confirmed influenza. Of these 43, the following percentages of subtypes were identified: influenza A/H1N1 (42%), seasonal A/H3 (33%), A/ without identified subtype (7%) and influenza B (19%). We identified no deaths in pregnant women from SARI.

**Table 2.** Characteristics of the pregnant population with severe acute respiratory infection, age, influenza virus and subtypes, incidence, deaths, and lethality of the historical case series, state of Ceará, Brazil - 2013 - 2018.

| **Historical series – Year** | **2013 - 2018** |
| --- | --- |
| **Severe acute respiratory infection reported** | 3297 |
| **Pregnant women (N / %)** | 145 (4) |
| **Age (median, variation)** | 25.95<br>(15-44) |
| **Severe acute respiratory infection with diagnosis (N / %)** | 134 |
| Influenza | 43 (32) |
| Other viruses / etiologic agents | 11 (8) |
| Unspecified | 80 (60) |
| **Incidence of severe acute respiratory infection due to influenza laboratory certificate (%)** | 32.1 |
| **Influenza subtypes:** | 43 (100) |
| *A H1N1 (N / %)* | 18 (42) |
| *A H1 / seasonal (N / %)* | - |
| *A H3 / seasonal (N / %)* | 14 (33) |
| *A without subtype (N / %)* | 3 (7) |
| *B (N / %)* | 8 (19) |
| **Deaths of pregnant women with severe acute respiratory infection (N / %)** | 3 |
| Deaths due to influenza | 0 (0) |
| Deaths due to other viruses / etiologic agents / unspecified | 3 (100) |
| **Lethality of severe acute respiratory infection by influenza laboratory certificate** |  |
| Lethality - influenza (%) | 0 |

**Source:** Secretary of Health of the State of Ceará; SINAM case report form for severe acute respiratory infection.

To better visualize the relationship between birth outcomes, SARI, and influenza we overlaid sets of data on the same plot. **Figure 4A** combines birth weight data with SARI data for 2018. We see birth weights fall as peak SARI and influenza season approaches. Birth weights peaked in the first week of the year then fell to their nadir by week 15, with a ~40 gram decline in average birth weight. Following cessation of the influenza vaccination campaign, SARI cases declined and average birth weights returned to a yearly average. **Figure 4B** compares average gestational scores with SARI data for 2018, revealing a pattern of lower average gestational scores early in the year and during influenza season, indicating a higher proportion of preterm births.

**Figure 4.**
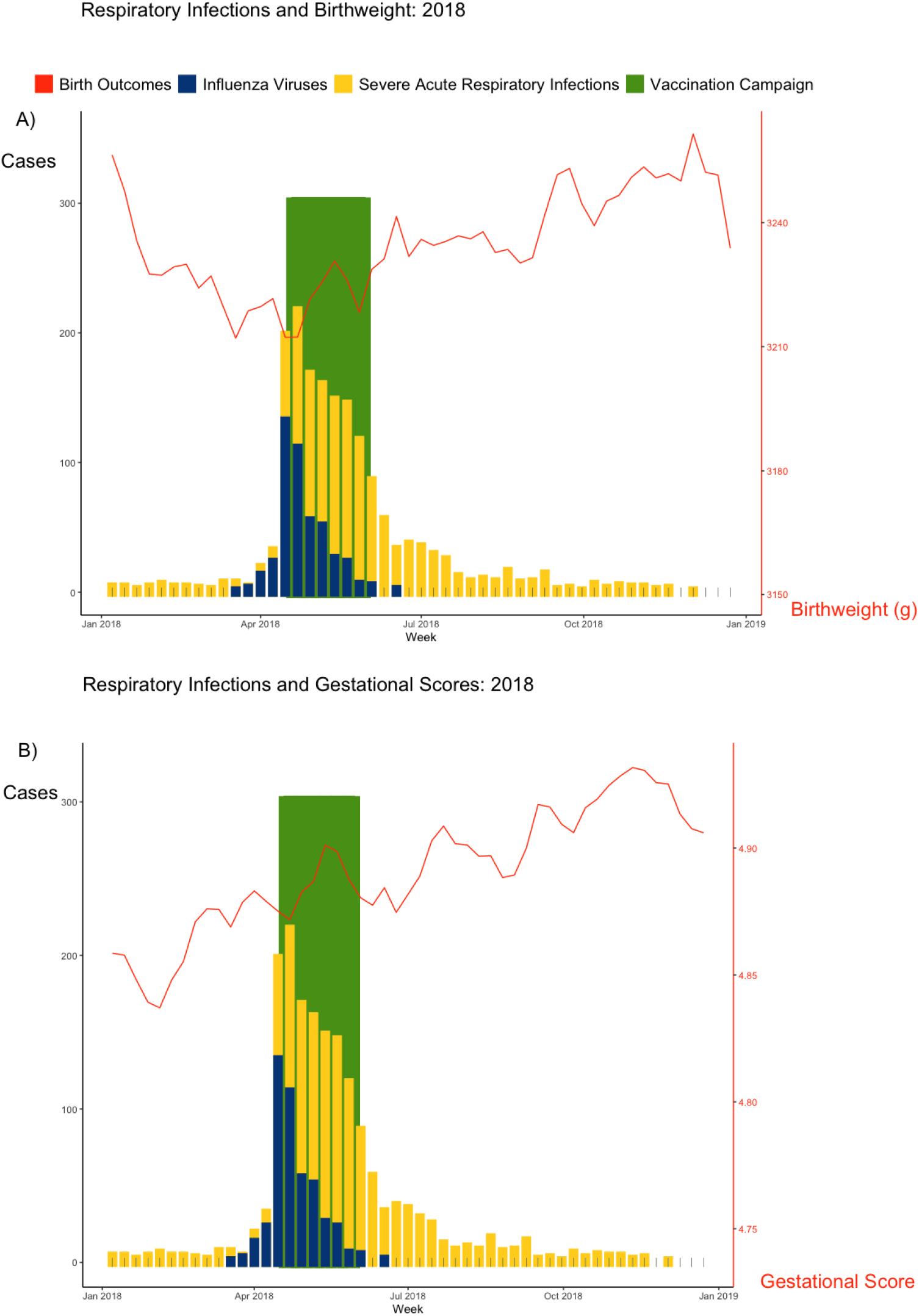
Influenza incidence, birth weights, and gestational length in Ceará, Brazil 2018. **(A)** Influenza incidence co-plotted with average weekly birth weights for 2018. Birth weight averages declined early in the calendar year, ahead of a spike in influenza cases at epidemiologic week 15 that coincided with an annual nadir in birth weights. Birth weights subsequently increased with the start of the vaccination campaign. By the time influenza cases decline, average birth weights reach the 2018 mean of 3220 grams. **(B)** Average gestational length oscillates throughout the year, remaining low early in the year and rising thereafter. Notably, the drop in influenza cases, the return to baseline, and subsequent increase in birth weights and gestational score occur following the vaccination campaign.

Annual periodicity of birth weights and gestational length from 2013 to 2018 are shown in **Figure 5**. On an annual basis, average birth weights oscillated by as much as 40 grams (or 1 to 2 percent of a child’s total birth weight). When we turned our lens towards gestational length, we found a similar pattern. 15·5% (8399/54311) of children born in February (a month associated with worse birth outcomes) were born preterm (<37 weeks). In contrast, 10·7% (6552/61067) of children born in October (a month associated with better birth outcomes) were born preterm. While much of the analysis above connects harmful effects of influenza in pregnancy on individual birth outcomes, the data in Figures 4 and 5 make the case that circannual oscillations in birth outcomes can be partially attributed to seasonal fluctuation and SARI and influenza in Ceará.

**Figure 5:**
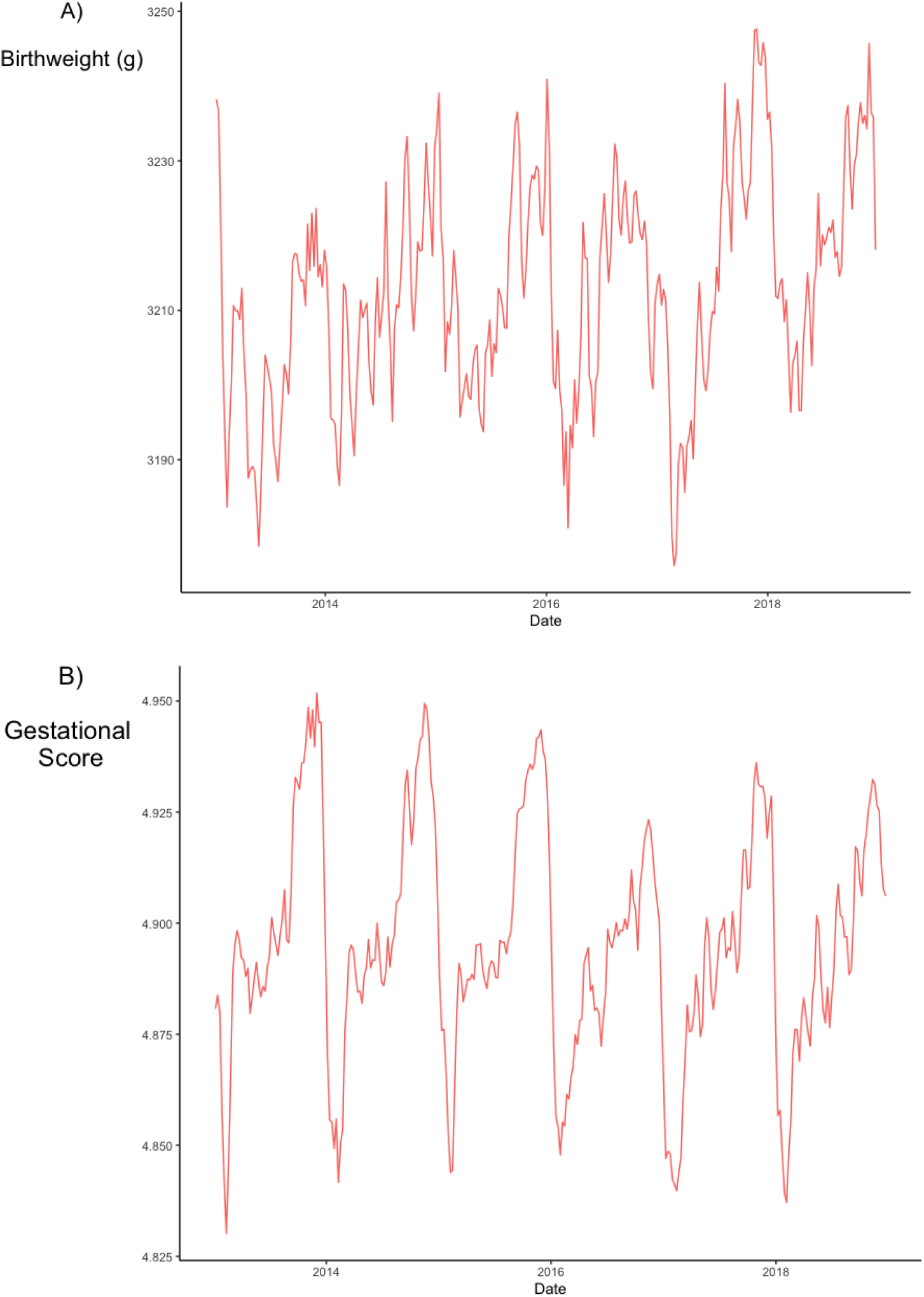
Circannual rhythmicity in birth weight and gestational age, Ceará, Brazil 2013-2018. Fluctuations in mean birth weight varied by as much as 2% of total weight at birth (70 gram) across seasons. For gestational lengths, we observed a repeating pattern which is characterized by a sharp decline in gestational length overlapping with influenza season. As the year progresses, the average gestational length increases before declining again.

Univariate analysis of cases (defined as children born to mothers with SARI) compared to controls (children of mothers who did not have a SARI) is presented in **Table 3**. Case children had significantly lower birth weights and higher risk of prematurity compared to controls. The 5-minute Apgar score was significantly lower in children born to mothers with SARI versus controls. The distribution of maternal education was significantly different between the two groups, with lower formal maternal education levels in cases. Mothers of controls had a significantly higher number of previous pregnancies, vaginal births and live births compared to mothers in the case group. The number of cesarean sections was higher in mothers of children in the case group compared to those in the control group. Cases had significantly shorter gestation times compared to controls. Cesarean deliveries and medical assistance were more frequent in cases versus controls (**Table 3**).

**Table 3.** Characterization of the population of liveborn children to pregnant women who had a severe acute respiratory infection (Cases) and children from pregnant women who did not report age-matched severe acute respiratory infections (Controls), in the historical series in the state of Ceará, Brazil, 2013 - 2018.

| **Variables** | **Total**<br>**N = 183** | **Cases**<br>**N = 61** | **Controls**<br>**N = 122** | **P values** |
| --- | --- | --- | --- | --- |
| **Male gender** (N /%) | 81 (45) | 24 (39) | 57 (47) | 0.202 |
| **Birth weight** (g; mean and standard deviation) | 3090.1 ± 665.42 | 2879.1 ± 783.57 | 3195.6 ± 572.61 | **0.019** |
| **Preterm Birth**(< 37 weeks gestation; N / %) | 26 (19) | 16 (27) | 10 (13) | **0.025** |
| **Apgar Index** (mean and standard deviation) |  |  |  |  |
| 1 minute | 8.0 ± 1.45 | 7.9 ± 1.66 | 8.0 ± 1.32 | 0.833 |
| 5 minute | 9.0 ± 0.98 | 8.9 ± 0.75 | 9.1 ± 1.08 | **0.004** |
| **Mother's age** (mean and standard deviation) | 28.3 ± 6.65 | 28.3 ± 6.69 | 28.3 ± 6.66 | - |
| **Education** (N /%) |  |  |  | **<0.001** |
| No schooling | 0 (0.0) | 0 (0.0) | 0 (0.0) |  |
| Elementary I (1st. 4th Grade) | 8 (4.6) | 1 (1.7) | 7 (6.0) |  |
| Elementary II (5th. 8th Grade) | 19 (10.9) | 1 (1.7) | 18 (15.5) |  |
| Medium (former 2nd. Degree) | 60 (34.5) | 14 (24.1) | 46 (39.7) |  |
| Incomplete higher | 73 (42.0) | 34 (58.6) | 39 (33.6) |  |
| Graduated | 14 (8.0) | 8 (13.8) | 6 (5.2) |  |
| **Previous pregnancies** (median and variation) |  |  |  |  |
| Number of previous pregnancies | 2, 0-16 | 1, 0-7 | 2, 0-16 | **0.014** |
| Number of vaginal deliveries | 2, 0-12 | 0, 0-4 | 2, 0-12 | **<0.001** |
| Number of cesarean sections | 0, 0-04 | 0, 0-4 | 0, 0-02 | **<0.001** |
| Number of live births | 2, 0-12 | 1, 0-7 | 2, 0-12 | **0.001** |
| Number of fetal losses / abortions | 0, 0-04 | 0, 0-2 | 0, 0-04 | 0.158 |
| **Current pregnancy** (mean and standard deviation) |  |  |  |  |
| Number of weeks of gestation | 37.8 ± 3.15 | 36.9 ± 3.72 | 38.4 ± 2.50 | **0.007** |
| Number of prenatal consultations | 6.9 ± 3.15 | 7.0 ± 2.55 | 6.7 ± 2.16 | 0.557 |
| Pregnancy month when prenatal care started | 2.9 ± 1.38 | 2.8 ± 1.22 | 3.1 ± 1.48 | 0.142 |
| **Type of pregnancy** (N /%) |  |  |  |  |
| Only | 181 (98.9) | 59 (96.7) | 122 (100) | 0.110 |
| Double | 2 (1.1) | 2 (3.3) | - |  |
| Triple or more | - | - | - |  |
| **Fetus presentation at delivery (N /%)** |  |  |  |  |
| Cephalic | 138 (96.5) | 54 (93.1) | 84 (98.8) | 0.089 |
| Pelvic or Podalic | 5 (3.5) | 4 (6.9) | 1 (1.2) |  |
| Transversal | - | - | - |  |
| **Was labor induced? (N /%)** |  |  |  |  |
| Yes | 5 (3.6) | 4 (6.9) | 1 (1.2) | 0.097 |
| No | 134 (96.4) | 54 (93.1) | 80 (98.8) |  |
| **Type of delivery (N /%)** |  |  |  |  |
| Vaginal | 128 (70.7) | 8 (13.3) | 120 (99.2) | <0.001 |
| Cesarean | 53 (29.3) | 52 (86.7) | 1 (0.8) |  |
| **Did Cesarean section occur before labor started? (N /%)** |  |  |  |  |
| Yes | 29 (76.3) | 29 (76.3) | - | - |
| No | 9 (23.7) | 9 (23.7) | - |  |
| **Birth attended by: (N /%)** |  |  |  |  |
| Doctor | 122 (84.1) | 59 (98.3) | 63 (74.1) | 0.001 |
| Obstetric nurse | 6 (4.1) | 1 (1.7) | 5 (5.9) |  |
| Midwife | 9 (6.2) | - | 9 (10.6) |  |
| Others | 8 (5.5) | - | 8 (9.4) |  |

Mann-Whitney test was used for variables whose distribution was not normal and Chi-square analysis was used for contingency.

We used multiple logistic regression to identify predictor variables independently associated with case status. Initially, we selected variables identified by univariate analysis (**Table 3**). Of eleven significant variables identified, we found five showing collinearity >40%. A logistic regression model was run with the remaining variables. Our sample size was adequate (N = 123 cases analyzed) to run the logistic regression (*17,19)*. The overall model fit showed a chi-square value of 23·135, 6 d.f., and *P* <0·001. The Cox & Snell, Nagelkerke test showed variance between 17·1% and 23·2% in explaining the model using these variables. Inclusion of predictor variables increased model accuracy from 61% to 68%. **Table 4** summarizes the unadjusted (OR) and adjusted (AOR) odds ratios for birth weight, preterm birth, maternal education, number of previous pregnancies, number of weeks of gestation, and birth attended by doctors associated with liveborn children to pregnant women who had SARI. Multiple logistic regression analysis (AOR) showed only birth weight (*P* = 0·030) and attendance of birth by a physician (*P* = 0·037) were significantly associated with case status (**Figure 6** and **Table 4**).

**Table 4.**
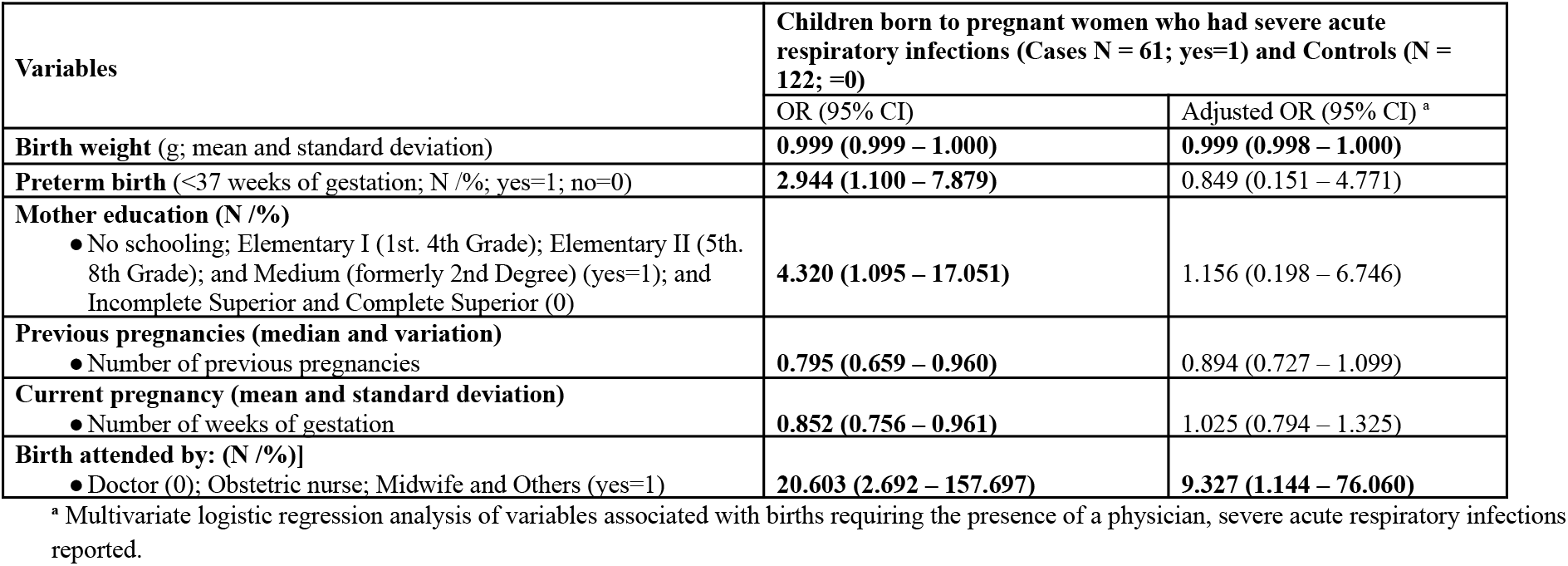
Odds ratio of children with birth weight, preterm, mother education, number of previous pregnancies, number of weeks of gestation and birth attended of pregnant women who had a severe acute respiratory infection reported in the historical series in the state of Ceará, Brazil, 2013 - 2018.

**Figure 6.**
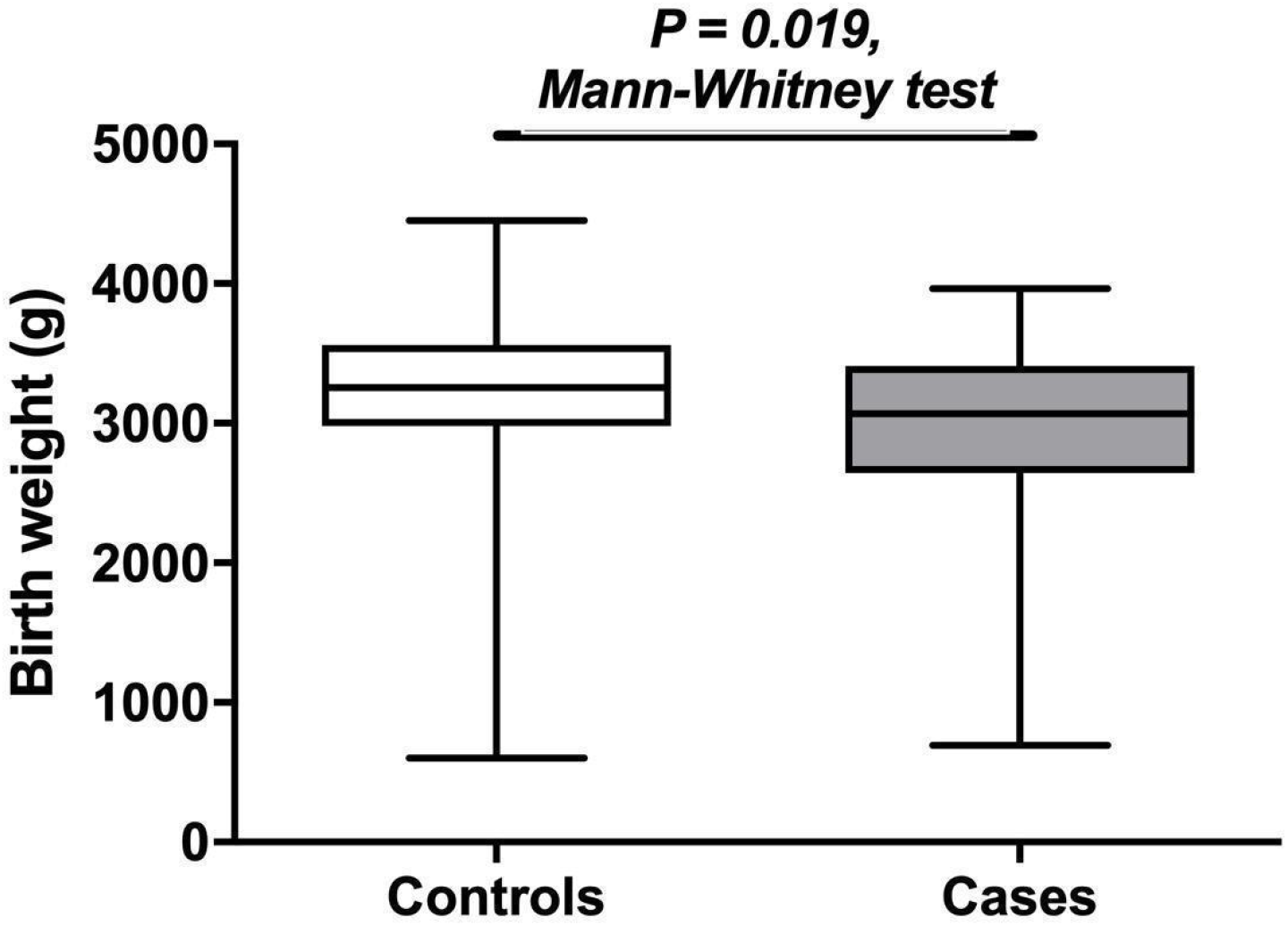
Reduced mean birth weights in children born to mothers with and without a severe acute respiratory infection during pregnancy. Box and whisker plot describes how children born to mothers who had a severe acute respiratory infection had significantly lower birth weights than control children born to mothers who had no documentation of a severe acute respiratory infection.

## Discussion

In this 5-year study of severe acute respiratory infection, influenza, and maternal-fetal outcomes in the Brazilian semiarid state of Ceará, we documented a total of 3,298 cases of severe acute respiratory infection between 2013 and 2018. Cases occurred predominantly in younger children—especially children under 6 months of age—followed by older adults and pregnant women. These data are consistent with data from prior literature showing higher rates of infection in younger populations but increased mortality in older adults (*20*).

Influenza A/H1N1 was the dominant strain in seasonal epidemic outbreaks, emphasizing the recirculation capacity of this strain and co-circulation with other seasonal strains (**Table 1** and **Figure 3A**). Lethality of the A / H1N1 strain in our time series was high. Moreover, we observed less lethality overall when seasonal A / H1 strain predominated in 2015. This data is consistent with prior literature showing excess mortality associated with A / H1N1 (*21*). The high lethality in our study may plausibly be attributed to the suboptimal timing of vaccination campaigns, which occurred during, or immediately following, peak influenza activity in Ceará. This is corroborated by the high percentage of vaccination coverage across the state, strongly suggesting administration of vaccines earlier in the year would reduce mortality and morbidity from influenza. Mistiming of influenza immunizations may also explain our finding of an unusually high disease burden in infants under 6 months. Immunization in this young age group is not recommended by WHO, hence immunization of pregnant mothers is especially important for passive immunity against influenza in the vulnerable first six months of life.

In this study, we assessed whether mothers with SARI were at higher risk of low birth weight or premature children. We found statewide correlations between peak influenza activity and nadirs in birth weight and gestational length. Further, we confirmed associations of maternal SARI with birth weight deficits and preterm birth in individual mother-infant dyads. These associations remained significant when adjusting for confounders by multiple logistic regression. The effect on birth weight is in accordance with recent data from two recently published studies, which demonstrated an association of SARI in pregnant women with or without influenza, with low-birth-weight outcomes (*6, 7*). The complications described here are underscored by the fact that medical assistance in labor was more prevalent in the group of children whose parents were affected by maternal SARI compared to the control group.

Our study aligns with earlier reports showing the importance of prevention and adjustment of influenza vaccine campaign schedules to avoid complications from influenza (*5, 22-25*). Previous studies show the importance of the first 1000 days of life in preventing childhood illnesses such as undernutrition, enteric infections and the subsequent risk of metabolic syndrome and cardiovascular diseases (*26,27*).

Neurocognitive, physical and educational deficits have been well documented for children exposed *in utero* or in the first months of life to influenza and other diseases such as enteric infections (*9-11,28*).

Our study had several limitations. First, we analyzed only cases of influenza associated with severe acute respiratory infection. Hence, cases of mild to moderate influenza were absent from the analysis. However, our statewide birth outcome analyses detected significant periodicity in birth weights and gestational length, with poorer outcomes coinciding with influenza season. This points to the possibility of larger scale effects of influenza on birth outcomes beyond SARI. Second, our nested observational descriptive study cannot infer a causal relationship between SARI in pregnancy and adverse birth outcomes. However, the associations were robust to logistic regression to adjust for several potential confounders. Additionally, while public hospitals are required to report cases of SARI to SINAM, private clinics can opt in to report. While many private clinics do report, most reported cases come from public institutions. As such, we cannot be certain we captured all cases of SARI across the state. Lastly, our results implicate imperfect timing influenza vaccination campaigns in adverse influenza outcomes in the Brazilian semiarid, but we did not model the extent to which earlier immunization or use of Northern versus Southern vaccine strains might mitigate these outcomes. Recent epidemiologic models suggest Ceará is the starting point for a wave of influenza transmission from the semiarid region to the South, hence immunization prior to peak activity in Ceará may have significant benefits for the region and wide swaths of the country (*4*). We did not account for infections with Zika virus as a potential confounder of our findings, however Zika incidence was zero from 2013 to 2016, with the caveat that testing not routinely performed during this time period. As shown by Ceará’s Secretary of Health (*29*), Zika incidence in Ceará was low at 5.6 and 0.2 per 100 thousand population in 2017 and 2018, respectively.

In conclusion, our results show suboptimal timing of influenza vaccination in a populous, semiarid Brazilian setting with high vaccine coverage contributes to an unacceptable--and partially preventable--burden of disease in pregnant women and young infants. In addition, we find annual periodicity in mean birth weights and rates of prematurity plausibly linked to seasonal influenza. Finally, we confirm a robust association of SARI during pregnancy and poor birth outcomes using an observational descriptive study design.

## Data Availability

The data that support the findings of this study are available from the corresponding author, JFQ, upon reasonable request.

## Acknowledgments

The authors thank the staff and data management team at Institute for Biomedicine, School of Medicine, Federal University of Ceará and the Epidemiological Surveillance Information System for Influenza, Ceará State Health Secretariat, Fortaleza, CE, Brazil for their contributions to this study. Dr. Mark Steinhoff was a major source of inspiration for these studies. We dedicate this manuscript to his memory.

## Author Bio

José Quirino da Silva Filho is an economist holding a Masters degree in Public Health, and is pursuing a PhD student in Medical Science. Additionally he works as a Datamanager at the UPC (Clinical Research Unit), IBIMED (Institute of Biomedicine) and INCT (National Institute of Biomedicine in the Brazilian Semi-Arid), Federal University of Ceará-Brazil. He served as a research fellow at the University of Virginia, VA, USA and participated in Data Center Coordination helping develop the data entry system of the international research network MAL-ED, funded by Bill and Melinda Gates Foundation.

## Author Contributions

**Conceptualization**: José Q Filho, Francisco S Junior, Thaisy BR Lima, Alberto M Soares, Jason A. Papin, Sean R Moore, Aldo A. M. Lima.

**Data curation**: José Q. Filho, Thaisy BR Lima, Alberto M. Soares, Vânia AF Viana, Jaqueline SV Burgoa, Aldo A. M. Lima.

**Formal analysis**: José Q. Filho, Francisco S Junior, Alberto M. Soares, Kunaal S Sarnaik, Gabriel A. Hanson, Aldo A. M. Lima.

**Investigation**: José Q. Filho, Francisco S Junior, Alberto M. Soares, Álvaro M Leite, Sean R Moore, Aldo A. M. Lima.

**Methodology**: José Q. Filho, Francisco S Junior, Alberto M. Soares, Vânia AF Viana, Jaqueline SV Burgoa, Álvaro M Leite, Sean R Moore, Aldo A. M. Lima.

**Resources**: José Q. Filho, Francisco S Junior, Alberto M. Soares, Sean R. Moore, Aldo A. M. Lima.

**Software**: José Q. Filho, Francisco S Junior, Alberto M. Soares, Aldo A. M. Lima.

**Supervision**: José Q. Filho, Francisco S Junior, Alberto M. Soares, Álvaro M Leite, Jason A. Papin, Sean R Moore, Aldo A. M. Lima.

**Validation**: José Q Filho, Francisco S Junior, Thaisy BR Lima, Vânia AF Viana, Jaqueline SV Burgoa, Alberto M Soares, Aldo A. M. Lima.

**Writing – original draft**: José Q Filho, Gabriel Hanson, Francisco S Junior, Alberto M Soares, Sean R Moore, Aldo A. M. Lima

**Writing - Revisions:** José Q Filho, Simone A Herron, Hunter L Newland, Kunaal S Sarnaik, Gabriel Hanson, Sean R Moore, Aldo A. M. Lima

## References

1. The top 10 causes of death [Internet]. Available from: https://www.who.int/news-room/fact-sheets/detail/the-top-10-causes-of-death

2. Wang X, Li Y, O’Brien KL, Madhi SA, Widdowson MA, Byass P, et al. Global burden of respiratory infections associated with seasonal influenza in children under 5 years in 2018: a systematic review and modelling study. Lancet Glob Heal. 2020 Apr 1;8(4):e497–510.

3. Simonsen L, Clarke MJ, Williamson GD, Stroup DF, Arden NH, Schonberger LB. The impact of influenza epidemics on mortality: Introducing a severity index. Am J Public Health. 1997;87(12):1944–50.

4. Katz MA, Gessner BD, Johnson J, et al. Incidence of influenza virus infection among pregnant women: A systematic review. BMC Pregnancy Childbirth. 2017;17(1). doi:10.1186/s12884-017-1333-5

5. Fell DB, Sprague AE, Liu N, Yasseen AS, Wen SW, Smith G, et al. H1N1 influenza vaccination during pregnancy and fetal and neonatal outcomes. Am J Public Health. 2012 Jun;102(6):e33.

6. Regan AK, Feldman BS, Azziz-Baumgartner E, Naleway AL, Williams J, Wyant BE, et al. An international cohort study of birth outcomes associated with hospitalized acute respiratory infection during pregnancy. J. Infect. 2020;81, 48–56.

7. Dawood FS, Kittikraisak W, Patel A, Hunt DR, Wesley MG, Thompson MG, et al. Incidence of influenza during pregnancy and association with pregnancy and perinatal outcomes in three middle-income countries: a multisite prospective longitudinal cohort study. Lancet Infect Dis. 2021;21, 97–106.

8. Mertz D, Geraci J, Winkup J, Gessner BD, Ortiz JR, Loeb M. Pregnancy as a risk factor for severe outcomes from influenza virus infection: A systematic review and meta-analysis of observational studies. Vol. 35, Vaccine. Elsevier Ltd; 2017. p. 521–8.

9. Black RE, Allen LH, Bhutta ZA, Caulfield LE, de Onis M, Ezzati M, et al. Maternal and child undernutrition: global and regional exposures and health consequences. Vol. 371, The Lancet. Elsevier; 2008. p. 243–60.

10. Victora CG, Adair L, Fall C, Hallal PC, Martorell R, Richter L, et al. Maternal and child undernutrition: consequences for adult health and human capital. Vol. 371, The Lancet. Lancet Publishing Group; 2008. p. 340–57.

11. Guerrant DI, Moore SR, Lima AAM, Patrick PD, Schorling JB, Guerrant RL. Association of early childhood diarrhea and cryptosporidiosis with impaired physical fitness and cognitive function four-seven years later in a poor urban community in northeast Brazil. Am J Trop Med Hyg. 1999;61(5):707–13.

12. MAL-ED Network Investigators. Early childhood cognitive development is affected by interactions among illness, diet, enteropathogens and the home environment: findings from the MAL-ED birth cohort study. BMJ Glob Health. 2018 Jul 23;3(4):e000752.

13. Takeda S, Hisano M, Komano J, Yamamoto H, Sago H, Yamaguchi K. Influenza vaccination during pregnancy and its usefulness to mothers and their young infants. J Infect Chemother. 2015;21(4):238–246. doi:10.1016/j.jiac.2015.01.015

14. Almeida A, Codeço C, Luz P. Seasonal dynamics of influenza in Brazil: The latitude effect. BMC Infect Dis. 2018 Dec 27;18(1).

15. Moura FEA, Perdigão ACB, Siqueira MM. Seasonality of influenza in the tropics: A distinct pattern in northeastern Brazil. Am J Trop Med Hyg. 2009 Jul 1;81(1):180–3.

16. Pallant J. SPSS survival manual : a step by step guide to data analysis using IBM SPSS. 352 p.

17. WHO information for the molecular detection of influenza viruses [Internet]. 2017. Available from: https://www.who.int/influenza/gisrs_laboratory/WHO_information_for_the_molecular_detection_of_influenza_viruses_20171023_Final.pdf

18. Monto AS, Gravenstein S, Elliott M, Colopy M, Schweinle J. Clinical signs and symptoms predicting influenza infection. Arch Intern Med. 2000 Nov 27;160(21):3243–7.

19. Kirkwood BR, Sterne JAC, Kirkwood BR. Essential medical statistics. Blackwell Science; 2003. 501 p.

20. Tabachnick BG, Fidell LS. Using multivariate statistics (6th ed.) New York: Harper and Row. 2012. 1024 p.

21. Simonsen L, Taylor R, Viboud C, Dushoff J, Miller M. US flu mortality estimates are based on solid science [2]. Vol. 332, British Medical Journal. BMJ; 2006. p. 177–8.

22. Glezen WP, Keitel WA, Taber LH, Piedra PA, Clover RD, Couch RB. Age distribution of patients with medically-attended illnesses caused by sequential variants of influenza A/H1N1: Comparison to age-specific infection rates, 1978-1989. Am J Epidemiol. 1991 Feb 1;133(3):296–304.

23. Bhat N, Wright JG, Broder KR, Murray EL, Greenberg ME, Glover MJ, et al. Influenza-Associated Deaths among Children in the United States, 2003–2004. N Engl J Med 2005 Dec 15;353(24):2559–67.

24. Omer SB, Goodman D, Steinhoff MC, Rochat R, Klugman KP, Stoll BJ, et al. Maternal influenza immunization and reduced likelihood of prematurity and small for gestational age births: A retrospective cohort study. PLoS Med. 2011 May;8(5).

25. Dodds L, MacDonald N, Scott J, Spencer A, Allen VM, McNeil S. The Association Between Influenza Vaccine in Pregnancy and Adverse Neonatal Outcomes. J Obstet Gynaecol Canada. 2012;34(8):714–20.

26. Giles ML, Krishnaswamy S, Macartney K, Cheng A. The safety of inactivated influenza vaccines in pregnancy for birth outcomes: a systematic review. Hum Vaccines Immunother. 2019 Mar 4;15(3):687–99.

27. Niehaus MD, Moore SR, Patrick PD, Derr LL, Lorntz B, Lima AA, et al. Early childhood diarrhea is associated with diminished cognitive function 4 to 7 years later in children in a northeast Brazilian shantytown. Am J Trop Med Hyg. 2002;66(5):590–3.

28. Lorntz B, Soares AM, Moore SR, Pinkerton R, Gansneder B, Bovbjerg VE, et al. Early childhood diarrhea predicts impaired school performance. Pediatr Infect Dis J. 2006;25(6):513–20.

29. Governo do Estado do Ceará. Secretaria de Saúde. (2018). Boletim Epidemiológico, Síndrome Congênita Associada à Infecção pelo vírus Zika. Available at: https://www.saude.ce.gov.br/wp-content/uploads/sites/9/2018/06/boletim_microcefalia_30_05_2018.pdf

